# Predictors of Post intensive care syndrome in pediatrics (PICS-p): A Systematic Review and Meta-Analysis protocol

**DOI:** 10.1101/2025.07.17.25331459

**Authors:** Sadaqat Ali, Laila Ladak, Qalab Abbas, Naveed Ur Rehman, Joseph C. Manning

## Abstract

**Introduction:** Post-Intensive Care Syndrome in Pediatrics (PICS-p) is being increasingly reported and investigated in children after a critical illness. This manifests as physical, cognitive, and mental problems that affect their quality of life. This systematic review aims to explore the risk factors leading to PICS-p to inform prevention and management strategies.

**Method and Analysis:** This systematic review and Meta-analysis will be carried out in accordance with the Preferred Reporting Items for Systematic Reviews (PRISMA) guidelines. The inclusion studies were quantitative cohort, case control, cross section and observational. Studies conducted in the last 2.5 decade (January 1, 2000 to January 1, 2025) involving pediatric population starting 1 month to 18 years of age, discharged from PICU. PubMed,Embase, Scopus and CINAHL databases will be used to search for the relevant studies. Boolean operations and Mesh terms will be used to ensure exhaustive literature coverage.

**Conclusion:** The review will provide an opportunity to know the risk factors non-modifiable (Age, sex, pre-existing health conditions and severity of illness) and modifiable (Delirium, sedation practice, PICU environment, Rehabilitation services and family support) that are associated with PICS-p and the findings can be used to make recommendations for targeted interventions to target interventions. Thus, by contributing toward overcoming this problem, the study is meant to improve the outcome for pediatric patients and families while decreasing future burdens due to long-lasting ailments. The outcomes are expected to shape clinical practices and policymaking in the direction of holistic care and recovery among survivors from a PICU.

**What is already known on this Topic?:** Despite growing recognition of post intensive care syndrome in pediatrics, there is no systematic review has been made regarding the factors that predispose to PICS-p in children with the age 1 month to 18 years, highlighting a huge knowledge gap.

**What this study hopes to add?:** The results of this systematic review identify risk factors linked to PICS-p and offer important new information for the future development of intervention programs for children’s risk factors modifiable and non-modifiable linked to PICS-p.

**How this study might affect research, practice or policy?:** This review is intended to present a body of evidence that will be useful in informing clinical practice and preventive efforts on preventive work for at risk children. The review will address how risk factor will help healthcare providers, researcher, policymakers, and caregivers in the long-term health and well-being of pediatric patients who will be discharged from PICUs.

## Introduction

Advancement in pediatric critical care medicine (PCCM) has reduced childhood mortality. As such, focus has been diverted to investigate the development of new morbidity following discharge from the Pediatric Intensive Care Unit (PICU) (1). Post-Intensive Care Syndrome in Pediatrics (PICS-p) is a framework that categorizes the decline in functioning or new onset impairments in physical, social, psychological and cognitive health domains following admission in the Pediatric Intensive Care Unit (PICU) (2). Childhood being a significant period of maturation and growth and that impact of survival can last days to decades (3).

Understanding the risk factors associated with PICS-p is crucial for planning effective management strategies for improving long-term outcomes in children. (4). Many studies have highlighted several risk factors of PICS-p, emphasizing the significance of clinical and demographic factors. For instance, the important one among these at the time of admission to the PICU was the severity of illness (5). Furthermore, length of stay in the PICU is a key risk factor for post-PICU syndrome, and studies reveal that longer PICU stays correlates with high rates of cognitive and psychological problems post-PICU hospitalization (6, 7). Approximately one in five children are diagnosed with PICS-p, defined by cognitive impairment when they spend four or more days in the PICU (6, 8).

The post-ICU pediatric population is likely to develop PICS-p, study showed that children with pre-existing neurological and neuromuscular conditions who rely on different levels of baseline-dependent care are more prone to develop PICS-p (9). Prolonged mechanical ventilation has also been implicated in higher risk (10). However, these results are not conclusive and may differ from study to study (6, 11). Long periods of bed rest during critical illness are associated with longer-term physical limitations post-discharge, which highlights the necessity of maintaining mobility and rehabilitation at the time of the ICU stay (12).

The psychological factors that contribute to PICS-p are important as well. The life of the child, as well as their family, can be greatly influenced by recovery outcomes (13, 14). Cognitive impairment is another critical driver of PICS-p, with reports suggesting attention, memory, and executive function problems among survivors (4, 15). Furthermore, benzodiazepine use as a common sedative and delirium risk factor has been identified as an additional contributor to the presence of both delirium and subsequently PICS in children, and in the PICU, the administration of narcotics and delusional memories has also been implicated as delaying recovery (16).

Understanding PICS-p risk factors will help in early interventions, enhanced patient outcomes, and the efficient use of healthcare resources. This review is intended to collect evidence that will be useful in informing clinical practice and preventive efforts on preventive work for at-risk children.

### Rationale and significance

PICS-p may lead to long-term functional disabilities, decreased quality of life, and high utilization of health resources (17). Identifying the risk factors of PICS-p will help in early interventions, enhanced patient outcomes, and the efficient use of healthcare resources. This review is intended to present a body of evidence that will be useful in informing clinical practice and preventive efforts on preventive work for at risk children. The review will address how risk factor will help healthcare providers, policymakers, and caregivers in the long-term health and well-being of pediatric patients who will be discharged from PICUs. Despite growing recognition of PICS-p, there is no systematic review has been made regarding the factors that predispose to PICS-p in children with the age 1 month to 18 years, highlighting a huge knowledge gap.

### Study objectives

To provide up-to-date evidence to identify risk factors associated with each component of post-intensive care syndrome in pediatric patients post PICU admission (PICS-p) in under 18 years of age.

### Review Questions

What are the risk factors for post-intensive care syndrome in pediatric (PICS-p) patients discharged from the PICU?

**PEO**

**P** (Population): Children aged 1 month to 18 years who have been discharged from a pediatric intensive care unit.

E (Exposure): PICU Exposure

**O** (Outcome): Risk Factors of developing Post-Intensive Care Syndrome in Pediatrics (PICS-p)

### Outcomes of study

- Identification of risk factors non-modifiable (e.g. Age, sex, pre-existing health conditions and severity of illness) and modifiable (e.g. Delirium, sedation practice, PICU environment, Rehabilitation services and family support) contributing to post-intensive care syndrome in pediatrics (PICS-p) among children I month to 18 years.

## Definitions

### PICS-p

PICS-p framework includes physical, cognitive, and psychological health issues that can occur in pediatric patients following discharge from PICU.

### Pediatric Patients

Population 1 month to18 years of age

### Discharged

Released from the PICU after getting treatment.

### PICU

Specialized unit for the treatment of critically ill children.

### Associated Risk Factors

PICS-p risk factors are clinical and demographic characteristics that are responsible for developing PICS-p in children who are getting discharged from the pediatric intensive care unit. Patient demographic characteristics include age, sex and socio-economic status. Clinical risk factors include medical and physiological factors, including pre-existing developmental disorders (Neurodevelopmental Delay, Cerebral Palsy, Autism Spectrum Disorder, Intellectual Disability, Attention-Deficit/Hyperactivity Disorder, Language and Speech Disorders, Learning Disabilities, Developmental Coordination Disorder), severity of the illness (Pediatric Risk of Mortality scale (PRISM) (PRISM III or IV), duration of mechanical ventilation, sedative exposure, sepsis or sever sepsis, Multiple Organ Dysfunction Syndrome, length of PICU stay, any invasive procedures (Central Venous Catheter Insertion, thoracotomy, Arterial Line Insertion, Tracheostomy, Dialysis Catheter Placement, emergency laparotomy, craniotomy, Lumbar Puncture and ) performed during critical care and other worth considering risk factors are diagnosis, duration of illness, immobility, hypoxia, hypotension, abnormal glucose and delirium.

## Materials and Methods

The Preferred Reporting Items for Systematic Reviews (PRISMA) guidelines will be followed to conduct this systematic review and Meta-analysis.

### Inclusion Criteria

- Observational research, including case-control, cross-sectional and cohort studies.
- Studies available in English language only will be Included.
- Time period January 1,2000 to January 1,2025
- Age 1 month to 18 years.
- Both gender.

### Exclusion Criteria

- Case studies, editorials, protocols, qualitative designs, and other reviews.

### Search Strategy

PubMed, Embase, Scopus and the CINAHL data base will be used to search for the relevant papers. We will combine Mesh terms with Boolean operators (AND, OR) to broadly search the topic and a preliminary pilot search with a librarian will be used to validate the search strategy.

**Search terms and alternatives following PICO :(P + E+ O)**

Search terms and alternatives following PICO :( P + E+ O)
| PEO | Terms | Mesh Terms |
| --- | --- | --- |
| **Population** | Children 1month to 18 years, discharged from PICU | child OR pediatric OR paediatric OR adolescent OR teens |
| **Exposure** | PICU Exposure more than 24hours | Post Intensive Care Syndrome OR PICS OR Post-Intensive Care Syndrome OR intensive care OR critical illness OR critical illness |
| **Outcomes** | Risk Factors of Post-Intensive Care Syndrome in Pediatrics (PICS-P) | predictor OR risk factor OR prognostic factor OR associated factor OR determinan OR ventilate OR sepsis OR ARDS OR sedation OR illness severity OR "cognitive disability OR "cognitive impairment OR "mental disability OR mental impairment OR physical disability OR physical impairment OR social disorder OR PTSD OR posttraumatic stress disorder |

### Final search

Post Intensive Care Syndrome OR PICS OR Post-Intensive Care Syndrome OR intensive care OR critical illness OR critical illness AND (child OR pediatric OR paediatric OR adolescent OR teens) AND (predictor OR risk factor OR prognostic factor OR associated factor OR determinan OR ventilate OR sepsis OR ARDS OR sedation OR illness severity OR “cognitive disability OR “cognitive impairment OR “mental disability OR mental impairment OR physical disability OR physical impairment OR social disorder OR PTSD OR posttraumatic stress disorder).

### Study Screening and Selection

Following the final search, all related articles will be uploaded to Covidence software, and duplicate articles will be eliminated. Two independent reviewers (SA and LL) will screen abstracts and titles to determine eligibility based on the inclusion criteria established for this review. Disagreement between any two reviewers would be referred to the third reviewer (QA or NRS) for resolution if any occur. All studies that potentially meet the inclusion criteria will have their full-text articles retrieved. After studying the article, two independent reviewers will assess it for eligibility. We will settle any eventual disagreements through discussion or with the assistance of a third reviewer. We will exclude studies in the full text if they do not meet the inclusion criteria. The final systematic review report will describe the findings of the study selection and present them in a PRISMA flow diagram with a description (Appendix 1).

### Data extraction

According to the study objective, data extraction sheet (Appendix 4) will be used to extract compulsory information from selected studies (SA and LL). Additionally, following information like Study ID, Study Design, Sample Size, Age, PICU Diagnosis, PRISM/PIM Score, Length of PICU Stay, Mechanical Ventilation, Sedation Use, Sepsis, Muscle Weakness, Mobility Issues, Fatigue, Exercise Intolerance, Anxiety, Depression, PTSD, Sleep Disorders, Memory Deficits, Attention Problems, Executive Dysfunction, Learning Difficulties, Parental Stress, Family SES, Follow-up Access, Premorbid Conditions, Developmental Stage, Ventilation Duration, Delirium, Multi-organ Failure, Parental PTSD, Parental Depression, Parental Anxiety, Socioeconomic Status and Insurance Coverage will be extracted and variables depends on availability.

### Quality assessment

Two independent reviewers (SA and LL) will critically evaluate the studies methodological quality. A third reviewer (QA or NR) will resolve any disputes that may eventually arise between the reviewers. The relevant critical appraisal tool will be chosen for each methodological design. The checklist for non-randomize studies (Appendix 2 and 3), is the standard Newcastle - Ottawa quality assessment scale assessment tool that reviewers will employ. It was made to rate the quality of studies that weren’t random, and its structure, content, and ease of use were all aimed at making it easy to use quality ratings when figuring out what meta-analytic results mean (18.

### Results and Meta-Analysis

The results will be analyzed using statistical software RevMan 5.4 version. Will check effect size calculation to check the association between risk factors and PICS-P, odds ratio (OR) for binary outcomes like presence vs. absence of PICS-p, relative risk (RR) to compare exposed vs. unexposed groups for different variables and mean difference for continuous variables, e.g., length of stay, mechanical ventilation.

In Meta-Analysis, will do a heterogeneity assessment to evaluate the variability between included studies, I2 statistic to measure the percentage of total variation due to heterogeneity (low: <25%, moderate: 25-50%, high: >50%), Cochran’s Q Test to assess whether observed differences are due to chance (p < 0.05 indicates significant heterogeneity) and variance estimation among studies in a random-effects model.

The subgroup analysis to identify how different groups influence the association between risk factors and PICS-p. For instance, age groups, infants vs. older children. PICU Stay Duration, Mechanical Ventilation Duration, Severity of Illness and Sedative Exposure etc.

### Strengths and Limitations

The review will cover the years between 2000 and 2025 and study published in English language only. However, finding the best PICU practices regarding the efficacy of the PICS-p intervention for children that aims to address risk factors related to PICS-p will be one of the review’s strengths.

## Data Availability

All data produced are available online at https://www.crd.york.ac.uk/PROSPERO/view/CRD420251041441

## Conclusions

The results of this systematic review identify risk factors linked to PICS-p and offer important new information for the future development of intervention programs for children’s risk factors modifiable and non-modifiable linked to PICS-p.

## Author Contributions

SA and LL: Conceptualization, Methodology and Software data curation, QA, NR: Supervision and Visualization, JCM: Reviewing, Editing and Supervision.

## Institutional Review Board Statement

This systematic review protocol is registered with the International Prospective Register of Systematic Reviews (Prospero) and registration number is PROSPERO 2025 CRD420251041441.

## Informed Consent Statement

Not applicable.

## Data Availability Statement

Not applicable.

## Conflicts of Interest

Nil

## Funding

Nil

## Appendix 01: PRISMA 2020 flow diagram

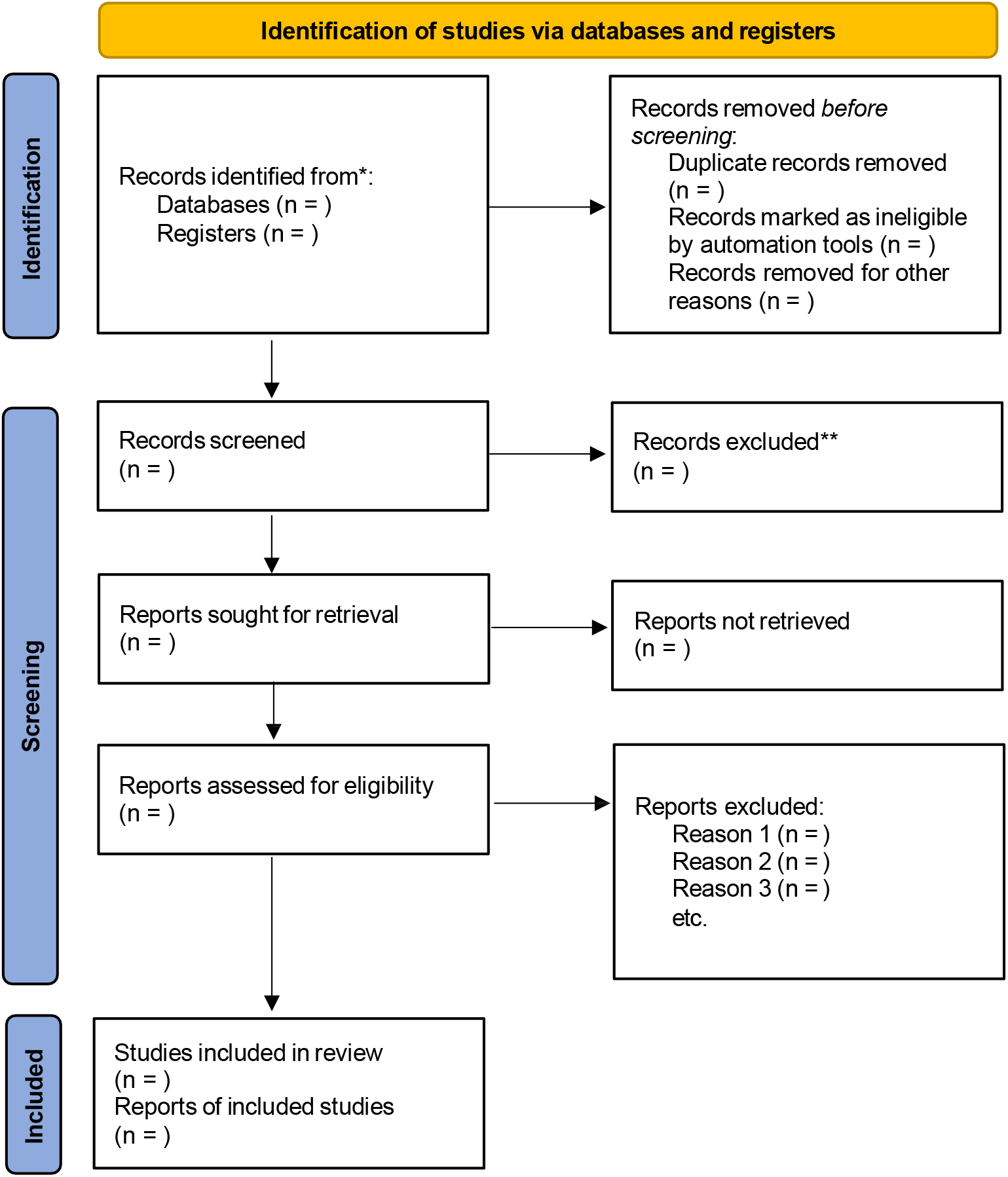

*Consider, if feasible to do so, reporting the number of records identified from each database or register searched (rather than the total number across all databases/registers).

**If automation tools were used, indicate how many records were excluded by a human and how many were excluded by automation tools.

Source: Page MJ, et al. BMJ 2021;372:n71. doi: 10.1136/bmj.n71.

This work is licensed under CC BY 4.0. To view a copy of this license, visit https://creativecommons.org/licenses/by/4.0

## Appendix 02: NEWCASTLE - OTTAWA QUALITY ASSESSMENT SCALE CASE CONTROL STUDIES

Note: A study can be awarded a maximum of one star for each numbered item within the Selection and Exposure categories. A maximum of two stars can be given for Comparability.

### Selection

1. Is the case definition adequate?
  a. Yes, with independent validation **\***
  b. Yes, e.g. record linkage or based on self-reports
  c. No description
2. Representativeness of the cases
  a. Consecutive or obviously representative series of cases *
  b. Potential for selection biases or not stated
3. Selection of Controls
  a. Community controls **\***
  b. Hospital controls
  c. No description
4. Definition of Controls
  a. No history of disease (endpoint) **\***
  b. No description of source

### Comparability

1. Comparability of cases and controls on the basis of the design or analysis
  a. Study controls for (Select the most important factor.) **\***
  b. Study controls for any additional factor **\*** (This criteria could be modified to indicate specific control for a second important factor.)

### Exposure

1. Ascertainment of exposure
  a. Secure record (e.g. surgical records) **\***
  b. Structured interview where blind to case/control status **\***
  c. Interview not blinded to case/control status
  d. Written self-report or medical record only
  e. No description
2. Same method of ascertainment for cases and controls
  a. Yes **\***
  b. No
3. Non-Response rate
  a. Same rate for both groups **\***
  b. Non respondents described
  c. Rate different and no designation

## Appendix 03: NEWCASTLE - OTTAWA QUALITY ASSESSMENT SCALE

**COHORT STUDIES**

Note: A study can be awarded a maximum of one star for each numbered item within the Selection and Outcome categories. A maximum of two stars can be given for Comparability

### Selection

1. Representativeness of the exposed cohort
  a. Truly representative of the average (describe) in the community **\***
  b. Somewhat representative of the average in the community **\***
  c. Selected group of users e.g. nurses, volunteers
  d. No description of the derivation of the cohort
2. Selection of the non-exposed cohort
  a. Drawn from the same community as the exposed cohort **\***
  b. Drawn from a different source
  c. No description of the derivation of the non-exposed cohort
3. Ascertainment of exposure
  a. Secure record (e.g. surgical records) **\***
  b. Structured interview **\***
  c. Written self-report
  d. No description
4. Demonstration that outcome of interest was not present at start of study
  a. Yes **\***
  b. No

### Comparability

1. Comparability of cohorts on the basis of the design or analysis
  a. Study controls for (select the most important factor) **\***
  b. Study controls for any additional factor **\*** (This criteria could be modified to indicate specific control for a second important factor.)

### Outcome

1. Assessment of outcome
  a. Independent blind assessment **\***
  b. Record linkage **\***
  c. Self-report
  d. No description
2. Was follow-up long enough for outcomes to occur
  a. Yes (select an adequate follow up period for outcome of interest) **\***
  b. No
3. Adequacy of follow up of cohorts
  a. Complete follow up - all subjects accounted for **\***
  b. Subjects lost to follow up unlikely to introduce bias - small number lost - > % (select an adequate %) follow up, or description provided of those lost)
  c. Follow up rate < % (select an adequate %) and no description of those lost
  d. No statement

## Notes

### Competing Interest Statement

The authors have declared no competing interest.

### Clinical Protocols

https://www.crd.york.ac.uk/PROSPERO/view/CRD420251041441

### Funding Statement

This study did not receive any funding

### Author Declarations

PubMed, Embase, Scopus and CINAHL databases.

